# Community Perceptions of Vaccination Among Influential Stakeholders: Qualitative Research in Rural India

**DOI:** 10.1101/2020.12.04.20244228

**Authors:** Baldeep K. Dhaliwal, Riti Chandrashekhar, Ananya Rattani, Rajeev Seth, Svea Closser, Anika Jain, David E. Bloom, Anita Shet

## Abstract

**Background:** In India and other low- and middle-income countries, multiple family and community members are influential in caregivers’ perceptions of vaccination. Existing literature indicates the primary caregiver, typically the mother, is instrumental in vaccine decision-making, but this may vary in contexts. We investigated the role of stakeholders in India who influence caregivers’ vaccination perceptions, as this is essential to developing strategies to promote vaccine acceptance and improve uptake.

**Methods:** This research was conducted in 2019 in Mewat District in Haryana, an area in India with extremely low vaccination coverage. We conducted six focus group discussions with 60 participants in the following categories: fathers of children under-5 years old, expectant mothers, mothers-in-law, community health workers, and community influencers such as locally elected officials and religious leaders.

**Results:** Our results highlighted four themes that influence vaccine uptake. First, while caregivers associated vaccination with reductions in specific diseases, they also noted that vaccination services brought broad health gains, including improved nutrition, antenatal guidance, and social support. Second, community health workers critically influenced, positively or negatively, caregivers’ vaccination perceptions. Third, community health workers faced gaps in their education such as limited training on vaccine side-effects, placing them at a disadvantage when dealing with families. Finally, we found that mothers-in-law, fathers, and religious leaders influence caregivers’ perceptions of vaccination.

**Conclusions:** Communication of broader benefits of vaccines and vaccination services by community health workers could be impactful in increasing vaccine acceptance. Vaccine uptake could potentially be improved by facilitating community health workers’ ownership over vaccine acceptance and uptake by involving them in the design and implementation of interventions to target mothers and mothers-in-law. A ‘bottom-up’ approach, leveraging community health workers’ knowledge to design interventions, and giving a voice to key members of the household and society beyond mothers alone, may sustain health improvement in low vaccine coverage areas.

## Background

Capturing nuanced perceptions of vaccination from groups that have decision-making power and influence within a community is essential to understanding and motivating vaccine acceptance and uptake. Although the Government of India has made efforts to improve vaccine uptake, according to data from the National Family Health Survey-4 (NFHS-4) conducted in 2015, only 13.1% of children under two years of age in Mewat, a district in Haryana, are reported to be fully immunized with bacilli Calmette-Guerin (BCG), measles, and three doses each of the polio and diphtheria-pertussis-tetanus (DPT) vaccine.^1^

Primary caregivers’ attitudes, beliefs, and acceptance of vaccination have been well studied in India, as well as in other low-and-middle-income settings globally.^2-4^ Community health workers (CHWs) also have significant influence on caregivers, and there have been several studies in India exploring CHW perceptions of vaccination, caregiver resistance to vaccinations, and the impact of vaccination campaigns on caregivers’ vaccination decisions.^5,6^ Studies that bring together select stakeholders-including CHWs, men, women, and health providers- to better understand perceptions of vaccinations have also been conducted in Indian states, but such studies consider these perspectives individually.^7,8^ Considering the perspectives of stakeholders in the context of each other is a critical omission of these studies, which we address herein.

Research has traditionally had an overemphasis on focusing on the role of caregivers, specifically mothers, in vaccine uptake and acceptance. While this perspective is essential, there has been limited attention to understanding the social context that influences this perspective. The perspectives of other influential groups that have decision-making power or influence over mothers’ vaccination decisions in India, including government-instituted community health workers, heads of households, and local community leaders, have been inadequately studied.

Gaining a well-rounded perspective on vaccine perceptions, particularly in low-resource settings like Mewat, is critical to understanding and overcoming barriers. Mothers in India have been found to have limited decision-making power in accessing healthcare and vaccination, and they depend on others, such as their mothers-in-law and husbands, to make those decisions.^9^ We brought together individuals that the community in Mewat considers influential, including village *sarpanches* (locally elected leaders), mothers-in-law, priests, and teachers, to understand how these stakeholders interact to impact caregivers’ vaccine decisions. Our expectation was that including this range of stakeholders would provide cross-cutting and holistic insights towards developing targeted interventions to promote vaccine acceptance and uptake in low-coverage areas.

### Background on India’s Community Health Worker Program

Vaccination services in India must reach more newborn children than in any other country, which requires a comprehensive health system and infrastructure.^10^ Within India there are three levels of community health workers established by the government: (1) an auxiliary nurse midwife (ANM) worker who provides vaccination services, (2) Anganwadi workers who primarily focus on childhood nutrition and preschool education and have a role in facilitating vaccination, and (3) accredited social health activists (ASHAs), subordinate to ANM workers, who critically contribute to the mobilization of rural mothers for vaccination. A brief table of their roles, and the health scheme in which they are nested is provided below (Table 1). These qualifications vary among states and districts based on local requirements.

**Table 1:**
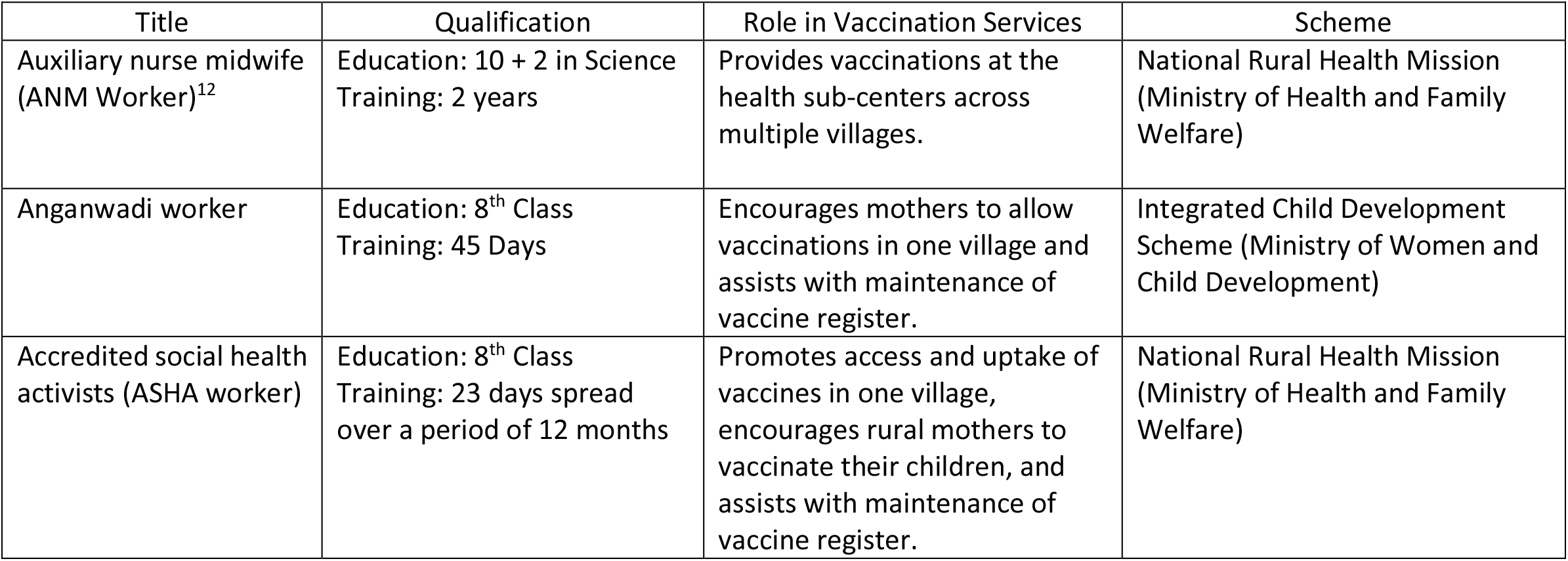
Community Health Workers in the Block Level^11^

| Title | Qualification | Role in Vaccination Services | Scheme |
| --- | --- | --- | --- |
| Auxiliary nurse midwife (ANM Worker)<sup>12</sup> | Education: 10 + 2 in Science<br>Training: 2 years | Provides vaccinations at the health sub-centers across multiple villages. | National Rural Health Mission (Ministry of Health and Family Welfare) |
| Anganwadi worker | Education: 8<sup>th</sup> Class<br>Training: 45 Days | Encourages mothers to allow vaccinations in one village and assists with maintenance of vaccine register. | Integrated Child Development Scheme (Ministry of Women and Child Development) |
| Accredited social health activists (ASHA worker) | Education: 8<sup>th</sup> Class<br>Training: 23 days spread over a period of 12 months | Promotes access and uptake of vaccines in one village, encourages rural mothers to vaccinate their children, and assists with maintenance of vaccine register. | National Rural Health Mission (Ministry of Health and Family Welfare) |

## Methods

### Study Procedures

We conducted a series of focus group discussions between October and November 2019 to understand perceptions of vaccinations from influential stakeholders in Mewat.

Recruitment took place in Patuka, Ghasera, Kurthala, Alduka, and Padheni villages in Mewat. These villages were selected because our team has strong relationships with Anganwadi centers, and the community members in these villages bring diverse perspectives. We recruited participants in the following groups: fathers of children under age 5, expectant mothers, mothers-in-law, Anganwadi and ASHA workers, and community influencers, such as the *sarpanch* (locally elected leader), religious leaders, and teachers. These groups were selected based on our teams’ deep knowledge of community members, and from preliminary conversations with caregivers to understand groups they find influential when making vaccination decisions.

We used lists of caregivers that Anganwadi and ASHA workers provided to sample participants for each group. Anganwadi and ASHA workers, as well as local study staff who are familiar with the community, helped identify and invite participants. Participants represented a range of ages, socioeconomic conditions, and family circumstances, including first-time pregnancies, caregivers who have lost children, and individuals who have worked as Anganwadi workers for twenty years. We made reminder calls and visits prior to the discussion, and we arranged focus groups in the local Anganwadi center. We trained a local team of research assistants to facilitate the focus groups. The local research assistants were well-known in the community, as they previously led research in Mewat and assisted with weekly mobile health clinics for village residents.

The number of participants per discussion group varied from 5 to 15, and the discussions lasted between 30 minutes and 1.5 hours. Questions and topics explored participant perceptions of benefits associated with vaccination and vaccine clinics, motivation for obtaining vaccinations for children, reasons associated with limited vaccine uptake and acceptance, and the role of the community and family in vaccination services. Each focus group was led in Hindi by one facilitator, while a second staff member took notes and assisted with group management. The focus groups were recorded digitally.

### Participants

We conducted six focus group discussions from October – November 2019 with a total of 60 participants (Table 2).

**Table 2:** Number of Focus Groups by Village and Participant Type (excluding member checking)

| Area and Participant Type | Village | Number of Focus Groups and Participants |
| --- | --- | --- |
| Anganwadi and ASHA workers | Ghasera | 2 (9F, 10F) |
| Fathers of children under 5 | Patuka | 1 (5M) |
| Community influencers | Alduka, Kurthala | 1 (9M and 1F) |
| Expectant mothers | Padheni | 1 (15F) |
| Mothers-in-law | Padheni | 1 (11F) |

### Analyses

The research team sent the audio recordings to an external agency for transcription and translation from Hindi into English. Our team conducted thematic analysis on the transcripts, and we developed a framework by inductively drafting a codebook of primary codes and sub-codes using grounded theory.^13^ Three individuals coded the transcripts separately and came together to compare findings and develop a codebook and framework for analysis. By using the framework and codebook to guide discussions, we established a consensus on data interpretation and gained an understanding of the prominent themes through weekly phone calls. To ensure a rich understanding of the language in the transcripts, we listened to the recordings and sought clarification on colloquialisms in the local language, Mewati, regularly. We organized quotes from the transcripts to understand the emergence of themes across multiple transcripts. Handwritten field notes from the focus group discussions were used to facilitate the interpretation of the data.

### Member Checking

We conducted synthesized member-checking with participants as part of data validation, where we verbally shared with participants the prominent themes that emerged from our analyses to assess if our results resonated with their experiences in Mewat.^14^ This process allowed us to validate our analyses and initial conclusions. Member-checking took place in July – August 2020, several months after completing data collection. To effectively enable engagement among a rural population in India, particularly during the COVID-19 pandemic, we chose to make individual phone calls to participants. We followed a novel five-step method of synthesized member-checking and made slight adjustments to adapt this method to our population (Figure 1).^15^

**Figure 1:**
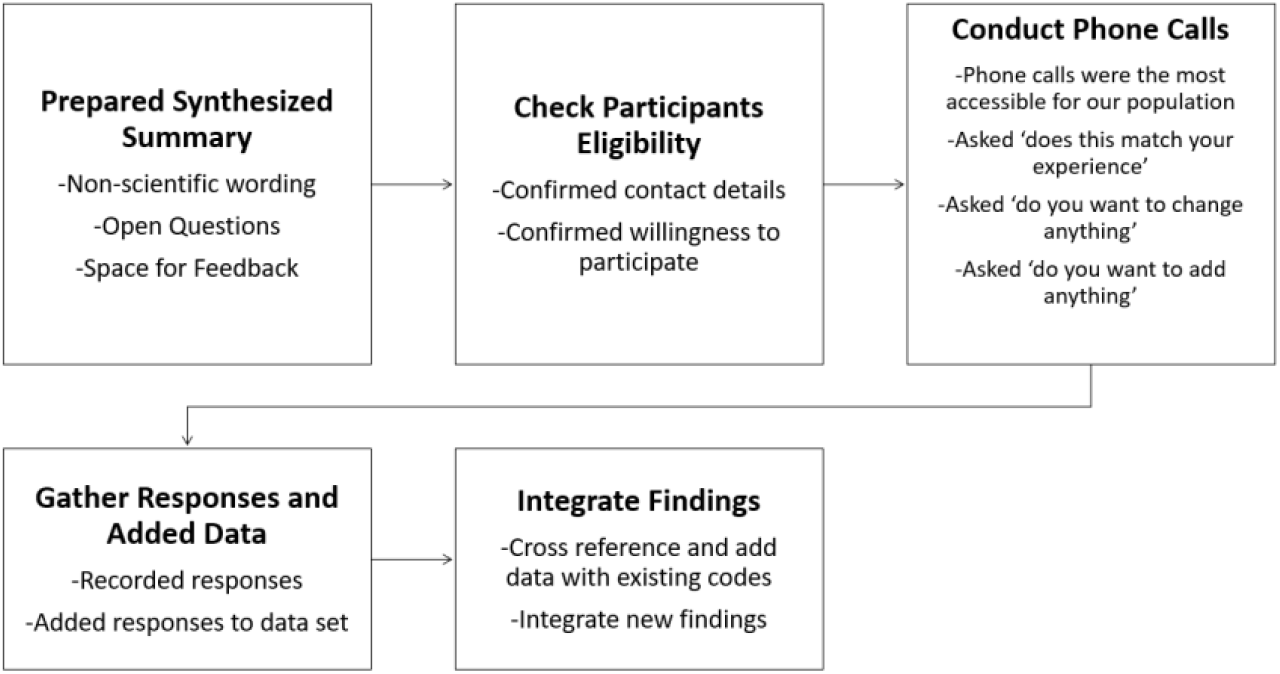
Processes Undertaken in Synthesized Member Checking

We completed member-checking with 21 focus group participants, and we had representation from each of our five participant groups. Overall, we found that the individuals who agreed to participate also agreed with the identified themes, which increased our confidence that our findings had captured participants’ experiences. In areas in which participants disagreed with our findings, our team discussed and re-evaluated our findings. Any discrepancies identified during member checking are explicitly stated.

### Ethical Considerations

We obtained ethical approval for the study from the Johns Hopkins Bloomberg School of Public Health Institutional Review Board (IRB) and the Ethics Committee of Bal Umang Drishya Sanstha (BUDS). Written consent was collected from all participants prior to engaging in data collection. All study participants’ identifiers were removed from the data transcripts to ensure anonymity of the study participants.

## Results

The themes that emerged in our results support four main messages. First, community members associated positive health outcomes and a broad set of benefits with vaccination and vaccination clinics. Second, Anganwadi and ASHA workers played a critical role in influencing, positively or negatively, caregivers’ perceptions of vaccination. Third, we found that despite their influential role, Anganwadi and ASHA workers still faced limitations that affect their empowerment and ownership over influencing vaccine acceptance and uptake. Fourth, we found that specific community and family members, beyond primary caregivers, are influential in caregivers’ vaccination decisions.

### 1) Vaccines are associated with positive health outcomes and broader benefits

#### Vaccine Preventable Diseases

Most participants believed that vaccination services contribute to disease reduction, and they also had specific knowledge of the diseases that vaccines protect against. Expectant mothers, mothers-in-laws, and community influencers discussed reductions in polio and measles in their communities as a result of vaccines. Expectant mothers and mothers-in-law were also aware that the BCG vaccine was given at birth, and in some cases could explain that it provided protection from tuberculosis. In the fathers’ group, however, there was less discussion and awareness of the diseases that were reduced as a result of vaccines, and they could only list polio as a vaccine-preventable disease. Anganwadi and AHSA workers were aware of these benefits, and they highlighted disease reduction as a motivating factor for promoting the use of vaccines.

> *“We go there [to family homes] thinking as if they are our own children. If we ignore them, then these children will not be healthy*.*” (ASHA Worker)*

Despite the differences in participants’ specific knowledge of the diseases that vaccines offer protection against, participants broadly highlighted disease reduction as a motivating factor to ensure children receive vaccination.

#### Broader Benefits of Services Provided with Vaccines

Participants from the expectant mothers, community influencers, and mothers-in-law focus groups highlighted a range of broader benefits that they associate with both vaccination clinics and Anganwadi and ASHA workers. Apart from disease reduction, they expressed that they received antenatal advice, nutrition guidance, and social benefits from vaccination visits to the Anganwadi center.

#### Antenatal Guidance

Expectant mothers related that when they receive their maternal vaccines, ASHA and ANM workers also provide them with antenatal care and advice that they would otherwise not receive.

> *“[They say] eat this food, it is good for health, take injections on time, check BP, go for test, all this information is given here*.*” (Expectant Mother)*

#### Nutrition Benefits

Participants highlighted that when they receive vaccines, they also receive information on the types of food they should, or should not, eat during their pregnancy, as well as the types of food they should be providing to their children.

> *“They tell us to eat green vegetables, taking food on time. [We] all come over here [Anganwadi center], and they tell us everything over here*.*” (Expectant Mother)*
>
> *“[We tell] that only mother’s milk is to be fed for six months. After six months child is to be given khichdi, porridge*.*” (ASHA Worker)*

#### Social Benefits

Participants also shared that they enjoyed being able to meet and socialize with other members of the community at the vaccination clinics.

> *“And the best benefit is that all are sitting together [to get vaccines]…Yes, the biggest benefit is that all are together, [with] the daughters, daughters in law*.*” (Mother-in-Law)*

Vaccination services that they receive at the Anganwadi centers also provide community members with an opportunity to learn from others in their discussions.

#### General Health Benefits

Participants also spoke broadly about the health benefits that they associate with vaccines.

> *“Nowadays there [are] lot of injections available, and each and every parent in the village gets their children immunized so that their children could get protected from all the diseases. Children are more healthy and fit now compared to previous times*.*” (Community Influencer, Sarpanch)*.

Participants had a sense that when children were fully vaccinated, they were generally healthier than children who were under-vaccinated or unvaccinated.

> *“All those whose children have got vaccinated they all are well and healthy. No child is a handicap*.*” (Mother-in-Law)*

### 2) The role of Anganwadi and ASHA workers is very important for vaccine uptake

The role of Anganwadi and ASHA workers was interwoven throughout the six focus group discussions, invoked when discussing trust, the benefits and side effects of vaccines, and when discussing who played an influential role in caregivers’ vaccine decisions. When we asked participants who they would turn to with their vaccination questions, or who provides them with vaccine knowledge, they invariably answered their Anganwadi or ASHA worker.

#### Vaccine Benefits

Providing information on, and access to, vaccination services was the most cited benefit of Anganwadi and ASHA workers. The population in Mewat, like other rural populations, finds it difficult to access vaccines due to their limited contact with the health system.

> *“We get them vaccinated here [Anganwadi center] only. If they get vaccinated here, then it’s good or else we ignore the vaccination*.*” (Father)*

Participants explained that ASHA workers provide the community with vaccine knowledge, access to vaccination services, and most importantly, they instill trust in the vaccine process.

> *“ASHA workers I don’t know [her] name they come to [our] home and tell us to come to center for getting vaccination done. If someone doesn’t come, still they come to remind. And in our village, they are doing very good work. They invite you so many times for vaccination*.*” (Mother-in-Law)*

#### Ability to Connect with Community

Anganwadi and ASHA workers shared that their deep level of knowledge, and familiarity with the community, enables them to lead targeted outreach in the community where vaccination campaigns have not reached, or in areas where they have not been successful. Anganwadi and ASHA workers are members of the community they serve, have frequent contact with caregivers, and complete regular home visits, positioning them to facilitate this outreach.

> *“Previously many people died in our village because of tetanus disease. Now we go from house to house where we know there [has been] a marriage, or where we know there are pregnant women. We search for them and now pregnant women are given vaccines*.*” (ASHA Worker)*

### 3) Community Health Workers have suboptimal ownership over vaccine acceptance and uptake

While members of the community highlighted the influential role of Anganwadi and ASHA workers, Anganwadi and ASHA workers shared challenges they face in promoting vaccine acceptance and uptake in Mewat. Gaps in education, limited training on side-effects, and negative perceptions of their roles were discussed as limitations, which all contribute to the overall challenge that Anganwadi and ASHA workers do not perceive ownership over vaccine uptake. While Anganwadi and ASHA workers expressed their desire to encourage caregivers to vaccinate their children, they also discussed the impediments to their actively influencing vaccine acceptance and uptake, as well as how these impediments can contribute to their decreased motivation.

#### Addressing Side-Effects

Participants discussed that there are many side effects to vaccines, ranging from swelling to fevers.

> *“Swelling occurs as well as he [child] gets fever. He cries so much that we can’t sleep at night, and in the morning, we have to get up early as well*.*” (Father)*

Anganwadi and ASHA workers explained that these side effects require constant attention from the primary caregiver, which is difficult for families to manage.

> *“Because it leads to fever, the child gets into problem and keeps crying for whole day. This means a full-time attendant is needed in house, this is the problem. If there is no one to look after child in home, then mother has to look after child as well as perform outdoor duties. Then it will create problem for her*.*” (ASHA Worker)*

Anganwadi and ASHA workers shared that they are neither equipped with medicines, nor are they trained to advise caregivers on the side effects children experience from vaccination. They rely on ANM workers to provide this information to caregivers. Although ANM workers have had extensive training on side effects and are best suited to provide this level of knowledge, they are also required to travel across villages, and they cannot consistently stay in one village long enough to comprehensively address these issues.

> *“I have such a case that the child in my area had a thigh vaccine. His thigh became so swollen that it still doesn’t resemble the other thigh. The ANM has not seen him, [even though] I sent her there. She has not visited till this date*… *For this reason, now none of the 20 families in that village come forward for vaccinations. Because of this [the ANM worker not addressing side effects], madam, I find it difficult to get children vaccinated*.*” (Anganwadi Worker)*

ASHA and Anganwadi workers are inevitably the ones who caregivers turn to, as they are consistently present in the community. When children experienced side effects that Anganwadi and ASHA workers cannot address, participants shared that this often led to negative perceptions of vaccines, frustrations with ASHA and Anganwadi workers, and ultimately caregiver resistance to vaccines.

#### Lack of Coordination and Delineation of Roles among CHWs

During focus groups with participants, it emerged that caregivers become frustrated with Anganwadi and ASHA workers for not addressing side effects, without realizing that ANM workers are better suited to address these issues, which contributes to caregivers’ larger anxieties about vaccines.

> *“Even if we give them vaccinations by convincing [caregivers of children], then we get [in] trouble because of swelling on the thighs of children. It causes swelling among children, then because of this they have fever for three days. Thereafter, problems arise in vaccination because some ladies say that our child becomes more ill [from vaccinations/side effects]*.*” (Anganwadi Worker)*.

This type of breakdown at the village level was discussed as contributing to distrust in the community, and Anganwadi and ASHA workers shared that this has the potential to jeopardize the success of vaccination programs in Mewat.

#### Limitations in Education and Training

Anganwadi and ASHA workers also shared that they themselves were not educated on vaccines until they underwent training for their positions.

> *“It is difficult to know about it ‘just like that’. When I started my work, then only I started to know more about vaccinations*.*” (ASHA Worker)*

Anganwadi and ASHA workers discussed how they often encounter caregivers in their community who raise concerns about rumors and side effects associated with vaccinating their children. Addressing these concerns is critical to empower community members to choose to vaccinate their child, but Anganwadi and ASHA workers are limited in their ability to respond to these specific concerns. These knowledge gaps were discussed as contributing to caregiver frustrations with CHWs and vaccination services.

#### Negative Perceptions of Anganwadi and ASHA Workers

Although many participants cited Anganwadi and ASHA workers as being their primary source of information on vaccines, many caregivers and community influencers still believe that they are not working to their full potential.

**“***Kids don’t get any medicine or anything else [from Anganwadi workers]*.*” (Father) “They [ASHA workers] take rounds, but sometimes only*.*” (Community Influencer, Teacher)*

These frustrations not only have the potential to limit their credibility in the community, they were also discussed as reasons for low vaccine uptake in Mewat. Anganwadi and ASHA workers also expressed that caregivers verbally, and sometimes physically, harass them due to these frustrations, limiting their ability and motivation to serve the community.

#### Community Rumors

Participants in the Anganwadi and ASHA worker and community influencer groups discussed that rumors consistently hinder their ability to increase support for vaccines throughout Mewat.

> *“There was a discussion a few days ago that people are getting injected for vasectomy*.*” (Community Influencer)*

The participants explained that rumors in Mewat predominantly link immunizations with sexuality and sterilization.

> *“They have said that girls will get [in] heat by this [Rota] vaccination, and they will run from home*.*” (Anganwadi Worker)*

These rumors have frightened and angered caregivers, leading to refusals of particular vaccines. These rumors, in conjunction with other factors, have made it difficult for Anganwadi and ASHA workers to motivate vaccine acceptance and uptake.

### 4) Non-caregivers are also influential in vaccine acceptance and uptake

Participants shared that members of the family and community, beyond primary caregivers, wield decision-making power and extensive influence on caregivers’ perceptions of, and decisions on, vaccination services.

#### Role of Women

Many of the participants explained that the responsibility for accessing vaccines for children lies with women, with mothers-in-law having the final decision-making power for vaccinating children.

> *“In home mostly there is the mother-in-law, so she tells her daughter in law to get the vaccination done and get it done for your child also. So, in home mostly mother in law can tell about all this because mostly gents are outside*.*” (Mother-in-Law)*

Although women are the primary vaccine decision makers, participants also discussed the lack of power younger women have in deciding to vaccinate their children.

#### Broader Familial Roles

Mothers-in-law discussed how they use their position as the matriarch to motivate their daughters-in-law to vaccinate their children. They also shared that they use their generational experiences of struggling to access vaccines when advocating for vaccine uptake for their grandchildren.

> *“No, we did not get for our children… Our grandchildren are having vaccinations now. Now, every child has a vaccine. We all make [the] children go to have the vaccine*.*” (Mother-in-Law)*

Anganwadi and ASHA workers are also aware of this dynamic, and they leverage the influence of the mothers-in-law when they are making home visits.

> *“I go to talk to the house elder, and tell her, ‘Mother, are you okay?’ I try to gain her trust because if she understands then she will send her daughter-in-law and all the children in the house. If the mother-in-law gets ready, then daughter-in-law automatically gets ready*.*” (Anganwadi Worker)*

During member checking, some Anganwadi and ASHA workers disagreed that the mother-in-law plays a large role in vaccine uptake in Mewat. They emphasized that vaccine decision-making is instead highly dependent on the family dynamics and relationship between the mother-in-law and daughter-in-law.

#### Role of Men and Gender Equity

While fathers stated that they were involved in discussions about vaccination at home, it was not considered appropriate for them to be further involved with the vaccination process; it is also considered inappropriate for Anganwadi and ASHA workers to engage directly with men. These cultural dynamics in Mewat create an environment in which there will inevitably be a gap in vaccine communications between men and Anganwadi and ASHA workers.

Men explained that they will avoid these settings, even allowing their children to miss routine vaccination services, for fear of creating an uncomfortable gender dynamic.

> *“We don’t go because most of the time we are outside of home because of work and if one man goes there, ladies will be uncomfortable, that’s why gents [don’t] go there*.*” (Father)*

Anganwadi and ASHA workers also discussed the role of men in the context of the harassment that women encounter from them for vaccinating their children. When fathers saw that their children had fevers and swelling, common side effects of vaccines, they did not allow their wives to continue accessing vaccines for their children. In some instances, participants discussed physical abuse emerging from this lack of understanding of side effects.

> *“In my area, there is one ironsmith, I gave [his baby] BCG then [the] husband didn’t say anything. When I gave Penta first then also he didn’t say anything. When I gave Penta second then the child got sore, so her husband came from somewhere and when he came he said, ‘why is the child getting fever? I am observing it since two days!’, so he started beating her [wife] up*.*” (Anganwadi Worker)*

These cultural dynamics create complex situations that can potentially lead to a gap in communication about side effects, and treatment for these side effects, between Anganwadi and ASHA workers, women, and men.

#### Broader Spiritual Influence

Throughout the community influencers and Anganwadi and ASHA worker focus groups, there were perceptions that certain groups in Mewat are less likely to get their children immunized. Many of the participants suggested that this was due to religious influences, but they did not provide specific details on their reasoning. In the mothers-in-law discussion, participants also emphasized that their faith plays a large role in the health of their grandchildren; this faith in a higher power was stronger than their faith in health providers.

> *“Over here [the] doctor keeps changing. It is not like [there is] only one doctor over here that we can trust. So, that’s why I said everything is done by God*.*” (Mother-in-Law)*

This lack of faith in health providers, and over-dependence on the divine, could potentially be indicative of community frustrations with doctors who only work in their area for short periods. Local community members seem to have limited trust in these doctors, as they regard them as ‘outsiders’ with high-turnover rates, further solidifying their trust in a higher power.

## Discussion

We identified several key messages from our focus group discussions with influential stakeholders in Mewat. First, community members are generally aware that vaccination leads to reductions in vaccine-preventable diseases, but they also tend to associate vaccination services with broader health benefits, such as nutrition and antenatal guidance. Second, Anganwadi and ASHA workers are familiar to the community, and they play a critical role, whether positive or negative, in impacting caregivers’ perceptions of vaccination. Third, Anganwadi and ASHA workers face challenges as a result of their training, rumors in the community, and negative views from members of the community; these factors hinder Anganwadi and ASHA workers from perceiving ownership over vaccine acceptance, and ultimately limit their ability to motivate vaccine acceptance and uptake in Mewat. Last, community members beyond primary caregivers play an influential role in caregivers’ perceptions of vaccines and, ultimately, vaccine uptake and acceptance in the community.

Our data provide a unique addition to a small, but growing, collection of literature on the benefits of vaccination above and beyond protection of the vaccinated children from infection by the target pathogen.^16-18^ This body of literature has suggested that vaccine effectiveness alone does not fully explain all benefits of vaccination. We saw, similarly, that community members perceive some non-disease-specific benefits of vaccination services and clinics, including antenatal, nutrition, and social benefits. Our data points to the importance of effectively engaging Anganwadi and ASHA workers in communicating with caregivers about vaccination services, including communicating these broad benefits of vaccination and vaccination services. By ensuring that the Anganwadi and ASHA workers are trained to communicate additional benefits about vaccination, we can create targeted frameworks and pathways where Anganwadi and ASHA workers highlight these broad benefits. Involving a range of influential stakeholders in sharing these messages could also be successful in driving vaccine acceptance and uptake in Mewat.

Caregivers are also greatly impacted by their interactions with Anganwadi and ASHA workers in Mewat. Many of our participants felt that Anganwadi and ASHA workers provided access to vaccines, shared broad health knowledge that they would not otherwise have access to, and worked to identify members of the community who may be unvaccinated or under-vaccinated. However, interactions in which caregivers felt that they were not provided with adequate treatment or information were discussed as having far-reaching detrimental impacts on the community. These results highlight the weight that Anganwadi and ASHA workers are given in the community, and the need to facilitate and empower their ownership over vaccine acceptance and uptake. Anganwadi and ASHA workers can be very influential in caregivers’ vaccine perceptions in Mewat and are in a unique position to engage both community members and clinical providers. Although Anganwadi and ASHA workers may be well-positioned to develop and maintain strong relationships with community members, previous research in India has indicated they are not provided with sufficient resources or training, limiting their ability to facilitate change.^19-21^ Our research supplements this literature, but further suggests that Anganwadi and ASHA workers do not perceive ownership over vaccine acceptance and uptake.

Enhancing Anganwadi and ASHA workers’ ownership over vaccine acceptance may facilitate a sense of responsibility towards health services, which may ultimately improve care for the community. Previous research in India also supports this insight. Involving CHWs in interventions, even without taking specific steps to facilitate ownership, have led to an increase in the proportion of fully vaccinated children, with a mean increase of 27% in children’s coverage rates.^11^ Interventions to motivate vaccine acceptance through community ownership have been limited in rural India, and among these limited approaches, researchers have not relied on the deep community knowledge that Anganwadi and ASHA workers posess.^22^ Additional research from Zambia has suggested that focusing on community ownership, without involving and listening to health workers, can limit success.^23^ Involving health workers closely, paying attention to their fears, and addressing obstacles they face, have been suggested as critical elements of facilitating ownership.^23^ Our research supports this, as Anganwadi and ASHA workers shared that the limitations they face in the community have contributed to reductions in their ability and motivation to drive vaccine uptake and acceptance in the community. By engaging Anganwadi and ASHA workers and community members to develop and lead interventions to increase vaccine acceptance, we aim to go beyond merely engaging the community by laying the groundwork for increasing ownership. One strategy to involve them in this process could revolve around using human-centered design (HCD). HCD is based on the idea that the individuals who are best suited to design and implement creative, context-specific, interventions are the individuals who provide and advocate for vaccination services in the community. HCD has been used to drive innovation in healthcare and other industries, but it has been underexplored in improving vaccine uptake.^24^ Ensuring Anganwadi and ASHA workers have the resources, strategies, and skills necessary to design and sustain these changes will empower them and facilitate long-term success.

Anganwadi and ASHA workers discussed their strategies to motivate vaccine uptake in the community by connecting with mothers-in-law, an influential group. While our data highlight the importance of leveraging the influence of the mothers-in-law, our data also indicate the importance of targeting outreach to influential community members and family members beyond women. Research has indicated that community leaders and local government officials are influential in facilitating HPV vaccine uptake in low- and middle-income settings.^25,26^ Our research supplements these findings, but also highlights the power of additional groups. Fathers were described as having the power to prevent their wives from vaccinating their children; religious beliefs and religious leaders were also cited as carrying more weight in the community than local health providers. These groups have decision-making power and influence over primary caregivers, yet they are not fully engaged in vaccine acceptance and uptake, as it is not culturally appropriate for women to lead outreach strategies to men. We may see an increase in vaccine acceptance and uptake in Mewat by involving male health workers to complete targeted outreach to these additional influential groups. This strategy could be instrumental in ensuring sustained improvement of health outcomes in Mewat.

Building vaccine understanding and trust requires both a bottom-up and top-down approach. In order to address and improve vaccine acceptance among communities, top-down guidelines and implementation from the government as well as bottom-up action from groups at the local level are critical for equity-oriented policies.^27^ This study illuminates the nature, challenges, and importance of the bottom-up perspective, as well as strategies for advancing community-level acceptance and support. Perspectives, impediments, and actions from local groups can provide governments with guidance on addressing the complexity of developing responses to vaccine uptake. By basing responses on strategies to create a health-promoting environment, governments could facilitate sustained health outcomes at the local level.

This study is limited by several factors. Our study sample was obtained from Anganwadi registrars, whereas recruitment from other sites, such as religious sites, might have allowed us to reach a wider population. Additionally, participants came from five villages, which might not be representative of perspectives throughout Mewat. Another limitation is a possible volunteer bias, as participants who have extreme, whether positive or negative, feelings about vaccination services might have been more willing to participate in the focus groups and during member-checking. Additionally, participants who chose to participate in our focus groups might have higher levels of awareness and knowledge of vaccines, creating the possibility that our participants might have more vaccine knowledge than the rest of the Mewat community. Finally, we did not include ANM workers in the focus groups, as we did not want to create an uncomfortable environment for the ASHA workers whom they supervise. However, they bring an essential perspective that we were not able to capture.

## Conclusion

Research and interventions have traditionally over-focused on understanding and targeting the role of the women, particularly the mother, in vaccine decisions. As we saw in our research, many younger mothers in low-income settings lack decision-making power. It is essential to think broadly, beyond the role of the mother alone, and design strategies that target community members, including men and religious leaders, who have influence and decision-making power in the community. Furthermore, by facilitating community health workers’ ownership of vaccine uptake to address these gaps in approaches, we can improve vaccine delivery, bring agency to communities, implement lasting strategies for vaccine acceptance, and overcome context-specific barriers in areas of low vaccine coverage.

## Data Availability

Data sharing is not applicable to this article as no datasets were generated or analyzed during the current study. The focus group transcripts analyzed during the current study are available from the corresponding author upon reasonable request.

## Abbreviations

(BCG): Bacilli Calmette-Guerin
(DPT): diphtheria-pertussis-tetanus
(CHW): Community Health Worker
(ANM): Auxiliary Nurse Midwife
(ASHA): Accredited Social Health Activists

